# Medical Students’ Attitudes toward AI in Medicine and their Expectations for Medical Education

**DOI:** 10.1101/2023.07.19.23292877

**Authors:** Joachim Kimmerle, Jasmin Timm, Teresa Festl-Wietek, Ulrike Cress, Anne Herrmann-Werner

## Abstract

**Objectives:** Artificial intelligence (AI) is used in a variety of contexts in medicine. This involves the use of algorithms and software that analyze digital information to make diagnoses and suggest adapted therapies. It is unclear, however, what medical students know about AI in medicine, how they evaluate its application, and what they expect from their medical training accordingly. In the study presented here, we aimed at providing answers to these questions.

**Methods:** In this survey study, we asked medical students about their assessment of AI in medicine and recorded their ideas and suggestions for considering this topic in medical education. Fifty-eight medical students completed the survey.

**Results:** Almost all participants were aware of the use of AI in medicine and had an adequate understanding of it. They perceived AI in medicine to be reliable, trustworthy, and technically competent, but did not have much faith in it. They considered AI in medicine to be rather intelligent but not anthropomorphic. Participants were interested in the opportunities of AI in the medical context and wanted to learn more about it. They indicated that basic AI knowledge should be taught in medical studies, in particular, knowledge about modes of operation, ethics, areas of application, reliability, and possible risks.

**Conclusions:** We discuss the implications of these findings for the curricular development in medical education. Medical students need to be equipped with the knowledge and skills to use AI effectively and ethically in their future practice. This includes understanding the limitations and potential biases of AI algorithms by teaching the sensible use of human oversight and continuous monitoring to catch errors in AI algorithms and ensure that final decisions are made by human clinicians.

## Background

Artificial intelligence (AI) is increasingly being used in the medical field. AI in medicine is an umbrella term that describes the use of algorithms and software that analyze data and digital information to make diagnoses and suggest therapies ^1-3^. AI plays a role in imaging diagnostics, for example, in the evaluation of CT scans or skin images and many more ^4,5^. Doctors can be supported by decision-support systems to diagnose diseases. Other fields of application for AI in medicine are drug development and the personalization of treatments ^4,6^. At the same time, however, not much is known about what prospective medical doctors know about AI and its application in medicine, how they assess this development, what they expect from their training in this respect, and what exactly they would like to see implemented in medical curricula.

## Materials and methods

In a survey study, we used an online questionnaire to ask medical students about their understanding and assessment of AI in medicine and recorded their suggestions for considering this topic in medical education. The questionnaire was advertised in November 2022 via the e-mail distribution list of the medical student council of a German university. Students were provided with a link to access the survey. They participated voluntarily and gave written informed consent. Participation took about 10 minutes and was not compensated.

The inclusion criteria for this survey study were that the participants were students of medicine or related subjects (medical technology, dentistry, neuroscience), that they completed the entire survey, and that they consented to the use of their data. Exclusion criteria were that participants were not students, were studying other subjects, did not completed the entire survey, or did not provide written informed consent.

In an online questionnaire, we asked medical students about their understanding and assessment of AI in medicine and recorded their suggestions for considering this topic in medical education. The questionnaire was advertised in November 2022 via the e-mail distribution list of the medical student council of a German university. Students were provided with a link to access the survey. They participated voluntarily and gave written informed consent. Participation took about 10 minutes and was not compensated.

First, the questionnaire asked whether participants were aware of AI in medicine (yes/no question) and inquired for their understanding of AI in general (two open questions: “What do you understand by AI?”, “What areas of application for AI in medicine can you imagine?”). Then we provided a short neutral definition of AI in medicine to ensure all participants had a basic comprehension of the term. This definition introduced AI in medicine exactly as in the background section of this article (i.e., algorithms to make diagnoses, suggest adapted therapies; relevant in imaging diagnostics, decision support systems, drug development, personalization of treatments).

After that, we asked for the perceived reliability of AI (5 items), its perceived technical competence (5 items), and faith (4 items) following Madsen and Gregor ^7^, as well as the perceived trustworthiness of AI in medicine ^8^ (12 items) on 5-point Likert Scales ranging from *1= do not agree at* all to *5= agree completely*. We also captured the perceived intelligence (5 items) and anthropomorphism (4 items) on semantic differential scales ranging from 1-5 ^9,10^. The selection of the specific scale format in each case followed the guidelines of the relevant research literature. Translations of all these items can be found in Appendix A. We also asked participants about their previous experience with AI, whether basic AI knowledge should be provided in university courses (yes/no question), and what specific aspects should be implemented in medical education (the choices were: technical basics, modes of operation, legal aspects, ethics, areas of application, potential future developments, classification of AI reliability, possible risks, and current AI systems; multiple responses possible). Finally, we asked them in an open question which aspects they perceived as problematic about the use of AI in medicine.

## Results

Eighty-four participants clicked on the link, but 17 dropped out before finishing the survey (dropout rate: 20.24%) and nine indicated not being medical students. The remaining 58 participants (35 females; mean age = 24.51 years, SD = 3.56 years) replied to all questions. The vast majority of participants (94.83%) indicated that they were aware that AI was used in medicine. They showed an adequate understanding of AI, referring in the first open question to machine learning (48.28%), algorithms (58.62%), and neural networks (8.62%) as the most relevant aspects (multiple responses possible; scores add up to more than 100%). As an application of AI in medicine, reference was made primarily to its use in diagnostics (86.21%) and surgeries (27.59%).

The participants perceived AI in medicine to be fairly reliable (M=3.30; SD=0.69), trustworthy (M=3.58; SD=0.71), and technically competent (M=3.26; SD=0.71), but they did not have much faith in it (M=2.34; SD=0.71). Moreover, they perceived AI in medicine to be rather intelligent (M=3.75; SD=0.66), but not anthropomorphic (M=1.99; SD=0.64).

The participants indicated only to a moderate extent that they already had experience with AI (M=2.85; SD=1.41; on a 5-point scale), learned about AI in an educational context (M=2.67; SD=1.47), or experienced AI in a medical context (M=2.69; SD=1.43). There was a very high level of agreement, however, when asked whether they were interested in the possibilities of AI in the medical context (M=4.52; SD=0.71), would like to learn more about AI (M=4.38; SD=0.83), and would like to see AI addressed more extensively in medical teaching (M=4.17; SD=0.92).

Fifty participants (86.21%) agreed that basic AI knowledge should be taught in medical studies. In particular, the participants supported the teaching of knowledge about modes of operation (77.59%), ethics (75.86%), areas of application (75.86%), reliability (94.83%), and possible risks (89.66%). There was less support for teaching technical basics (46.55%), legal aspects (46.55%), future developments (46.55%), and current AI systems (43.10%). Potential problems of AI in medicine that were pointed out by participants in an open question included ethical concerns (53.45%), lack of control (43.10%), and the potential lack of reliability of AI (34.48%; multiple responses possible; scores add up to more than 100%).

## Discussion

The findings of this study indicate that medical students are very interested in AI in medicine and want to learn more about it in medical school ^11^. The interest of medical students in this topic is not surprising given the rapid advances in AI and its potential to revolutionize healthcare. Given the potential of AI in medicine, it appears to be important for medical schools to incorporate AI education into their curricula. It is noteworthy that participants were particularly interested in ethical aspects of AI use and pointed out ethical concerns. Therefore, medical education should address these issues, for example, by teaching the challenges of data protection and privacy as the use of AI in medicine often requires access to highly sensitive patient health data. Other ethical aspects that should be considered are transparency and explainability (since AI systems can often make complex and opaque decisions) as well as discrimination and bias (as AI systems can reinforce discrimination due to insufficient or biased training data).

It is a limitation of this study that its participant pool consisted only of medical students from one German university, which may reduce the generalizability of our findings. This limitation should be addressed in future studies by expanding the participant pool to other medical schools. Moreover, one fifth of participants dropped out before finishing the survey; in future studies, offering compensation could possibly reduce this rate. It is noticeable that participants indicated rather low faith in AI. The items of this scale (see Appendix A) were mainly aimed at situations in which the suitability of the AI statements is unclear, so that these concrete item formulations could be responsible for this finding. Since this was a survey study in which no hypotheses were tested, no power calculation was made in advance.

## Conclusions

Medical students need to be equipped with the knowledge and skills to use AI effectively and ethically in their future practice. This includes understanding the limitations and potential biases of AI algorithms. Potential risks of AI in medicine could be addressed by teaching the sensible use of human oversight and continuous monitoring to catch errors or biases in AI algorithms and ensure that final decisions are made by human clinicians. By taking these steps, medical education can ensure that AI in medicine is used effectively and safely to improve patient outcomes.

## Data Availability

All data produced in the present study are available upon reasonable request to the authors.

## Acknowledgements

All of the participants made their data available voluntarily and data analysis was carried out on the basis of anonymized data. Participants gave written informed consent and were informed about privacy protection, their right to withdraw the data at any time without disadvantages, and about the general purpose of the study. The study was reviewed and approved by the Ethics Committee of the Leibniz-Institut fuer Wissensmedien (approval number: LEK 2022/048).

## Authors’ Contributions

All authors contributed to study design, data collection, and data analysis. JK primarily wrote the manuscript with edits and review by JT, TFW, UC, and AHW. All authors have read and agreed to the published version of the manuscript.

